# Performance of the iBox prognostication system in African American kidney transplant recipients: a multicenter study

**DOI:** 10.64898/2025.12.01.25341345

**Authors:** Agathe Truchot, Yannis Lombardi, Marc Raynaud, Olivier Aubert, Gillian Divard, Thibaut Thalamas, Brad Astor, Didier Mandelbrot, Sandesh Parajuli, Kenneth A. Newell, Babak Orandi, John J. Friedewald, Gaurav Gupta, Enver Akalin, Stanley C. Jordan, Arthur J. Matas, Miklos Z. Molnar, Junji Yamauchi, Katalin Fornadi, Andrew J. Bentall, Mark D. Stegall, Roslyn B. Mannon, Dorry L. Segev, Edmund Huang, William E. Fitzsimmons, Alexandre Loupy

## Abstract

The iBox is a validated prognostication system that predicts graft loss in kidney transplant recipients (KTRs), but its performance in the specific population of African American recipients has not been fully studied. We conducted a multicenter study on 3,588 KTRs from North America, including 866 (24.1%) African American KTRs, to assess the impact of race on the iBox’s performance in predicting graft loss. Performance metrics for the prediction of graft loss were similar for both African American and non-African American KTRs in terms of discrimination (C-index 0.81 [95% CI: 0.78-0.84] and 0.83 [95%CI: 0.81-0.85], respectively, p=0.25), and calibration (observed/expected ratio 1.13 [95%CI: 1.03-1.24] and 1.03 [95%CI: 0.95-1.11], respectively, p=0.13). No significant interaction between iBox score values and race was found in multivariate analysis stratified by transplant center (p=0.80 for the interaction term). Results were consistent regardless of the equation used to estimate glomerular filtration rate (Kidney Recipient Specific, CKD-EPI, or MDRD). In conclusion, the iBox prognostication system accurately predicts graft loss up to 7 years post-risk evaluation in African American KTRs and confirms its relevance as a surrogate endpoint in this population.

## INTRODUCTION

Improving long-term graft survival still remains one of the key unmet needs after kidney transplantation^1^. This challenge is exacerbated by the global organ shortage, which places a significant burden on healthcare systems worldwide, with demand far exceeding the available supply^2^. Recent advances regarding post-transplant management of kidney transplant recipients (KTRs) have mainly concerned diagnostic and prognostic tools, with a notable stagnation regarding therapeutics, the last approval regarding immunosuppressive therapies being belatacept in 2011^3^. These advancements, regarding patient monitoring and rejection diagnosis, have not been sufficient to overcome this issue of long-term graft survival, and new therapeutics are required to obtain an improvement in graft survival.

The integrative Box (iBox) prognostication system (NCT03474003) was specifically designed to address these limitations^4^. It has been shown that the iBox could serve as a surrogate endpoint in clinical trials^5^, and could hence fast-track the development and approval of pharmaceutical agents. This was acknowledged by the European Medicines Agency, which granted the iBox the qualification of being a “secondary endpoint prognostic for death-censored allograft loss (allograft failure)”^6^. This qualification allows it to be used in clinical trials to support the evaluation of novel immunosuppressive therapy applications^7,8^.

More recently, the US Food and Drug Administration (FDA) has accepted the Qualification Plan for the iBox prognostication system, as a reasonably like coprimary surrogate efficacy endpoint^9^. A crucial point to address, especially in the context of this qualification process, is the algorithm’s fairness^10,11^. A fair algorithm can be defined as one not “creating or exacerbating any disparities, in either provision of healthcare or subsequent healthcare outcomes”^10,11^. In the context of a clinical trial, fairness issues regarding outcome adjudication, i.e., systematic differences between ethnic groups in how the outcome performs, might lead to the wrongful restriction of a drug’s use in specific populations.

This aspect is particularly important for African American kidney transplant recipients, for whom it has been shown that disparities exist regarding access to and outcomes after kidney transplantation^12–14^. It has also been suggested that, in this specific population, the routine use of algorithms whose fairness was not properly assessed could have harmful impact on patients’ outcomes^15^. Additionally, the FDA strives to ensure the generalizability of results of new therapies across the target population; here, the general population of kidney transplant recipients. We hypothesize that the iBox prognostication system predicts outcomes similarly in African American and non-African American kidney transplant recipients.

The goal of this study was therefore to evaluate the performance of the iBox prognostication system in African Americans, using a large and multicentric cohort of North American kidney transplant recipients.

## METHODS

### Source and study population

The source population for this evaluation study comprised all kidney transplant recipients who were over 18 years of age at the time of transplantation and who underwent transplantation between: 2002 and 2017 at the University of Wisconsin Health University Hospital (Madison, WI, USA); 2004 and 2014 at the Johns Hopkins Medical Institute (Baltimore, MD, USA); 2009 and 2017 at the Northwestern Memorial Hospital (Chicago, IL, USA); 2010 and 2014 at the Montefiore Medical Center (New York, NY, USA); 2012 and 2016 at the Cedars Sinai Medical Center (Los Angeles, CA, USA); 2021 and 2024 at the University of Utah Health (Salt Lake City, UT, USA); 2003 and 2008 at the Mayo Clinic (Rochester, MN, USA); 2002 and 2016 at the Virginia Commonwealth University School of Medicine (Richmond, VA, USA); and 2005 and 2011 at 7 US and Canadian centers participating in the Deterioration of Kidney Allograft Function (DeKAF) multicenter cohort study^16^.

In all centers, data, including self-declared recipients’ race, were collected as part of routine clinical practice and entered into the centers’ databases in compliance with local and national regulatory requirements. In the DeKAF study, data collection was performed according to the study protocol. All clinical decisions (e.g., modalities of immunosuppression, decision to perform a biopsy), were made according to local practices and to the discretion of the treating physician. The transplantation allocation system followed the rules of the US Organ Procurement and Transplantation System in US centers, and those defined in each province in Canadian centers. All datasets were sent anonymized to the Paris Transplant Group.

The study population was obtained from the source population and consisted of 3,588 patients with (i) available data regarding race, and (ii) at least one computable iBox score in the post-transplantation period, defined as at least one adequate allograft biopsy (as per Banff criteria) performed during follow-up as part of the standard of care, and available data at the time of biopsy for eGFR, urine protein-to-creatinine ratio (uPCR), and anti-HLA donor-specific antibodies (DSA). For patients with multiple transplantations during the study period, only the first transplantation was considered.

### Outcome

The outcome to be predicted was death-censored graft loss, defined as a patient’s definitive return to dialysis or pre-emptive kidney re-transplantation. In each center, patients were followed up from the date of iBox evaluation (date of risk evaluation) until graft loss, death, loss to follow-up before the end of the study, or the end of the study, whichever came first. For patients who were followed for more than 7 years, follow-up was right-censored at 7 years in order to align with the original iBox prediction horizon.

#### The iBox prognostication system

The iBox prognostication system was designed to predict long-term death-censored graft loss and integrates eight independent parameters: 1) time between the date of transplantation and the date of risk evaluation (years), 2) eGFR (ml/min/1.73 m²), 3) urine protein: creatinine ratio (UPCR) (g/g, log-transformed), 4) circulating anti-HLA donor-specific antibody mean fluorescence intensity (binary cutoff of 1,400), 5) transplant glomerulopathy (cg Banff score), 6) microcirculation inflammation (g+ptc Banff score), 7) interstitial fibrosis and tubular atrophy (IFTA Banff score), and 8) interstitial inflammation and tubulitis (i+t Banff score), scored according to the Banff international classification.

In the main analysis, eGFR was estimated using the 2021 CKD-EPI race-free creatinine-based equation^17^. In sensitivity analyses, we assessed the robustness of the main findings when the eGFR was estimated using the MDRD_186_ creatinine-based equation^18^ (as in the original iBox publication), and the Kidney Recipient Specific (KRS) race-free creatinine-based equation^19^.

The iBox was computed at the time of risk evaluation after transplantation, which was conducted at the time of allograft biopsy performed for clinical indication or as *per protocol*, according to the centers’ practices. At the time of risk evaluation, recipients underwent concomitant evaluation of eGFR and proteinuria, allograft biopsy (Banff lesion scores and diagnoses), and circulating anti-HLA antibodies.

#### Statistical analysis

The performance of the iBox can be evaluated at any time point post-risk evaluation up to 7 years. For each patient, a predicted probability of graft loss at the time point considered for the analysis was generated using the iBox. Importantly, no recalibration or re-derivation was performed (i.e., the baseline survival function, centering constants, and β coefficients used to generate predictions were those of the original iBox).

We assessed the performance of the iBox based on its discrimination (ability to differentiate between patients with and without the outcome), calibration (agreement between the observed outcomes and estimated risks from the model), overall fit (i.e., accounting for both discrimination and calibration), and clinical utility.

The discrimination was assessed with Harrell’s concordance index^20^ (ideal: 1). The linear predictors of the iBox model were used as one-dimensional summaries of relative risk predictions for concordance evaluation^21^. 95% confidence intervals were calculated as described in Therneau and Atkinson^22^.

Calibration was assessed in terms of calibration slope (ideal: 1), calibration-in-the-large (ideal: 0), and observed/expected ratio (ideal: 1). The calibration slope over the entire follow-up period was derived by regressing the prognostic index (PI) values of the iBox model^23^. The observed/expected ratio was obtained by dividing the observed outcome proportion given by the Kaplan-Meier estimator by the mean of the predicted risks. Calibration metrics are presented along normal-based 95% confidence intervals. Calibration plots at 3-, 5-, and 7-years post-risk evaluation were derived by using the corresponding predicted risks from the iBox model were divided into equally-sized groups and, for each group, the median was plotted against the observed event probability estimated by one minus the Kaplan-Meier estimator.

The overall fit was assessed with the Brier Score (ideal: 0), the Index of Predictive Accuracy^24^ (ideal: 100%), and Royston and Sauerbrei’s R^2^ ^25^(ideal: 1). The Brier Score was calculated using inverse probability of censoring weighting, as defined by Gerds *et al*^26^. The Index of Predictive Accuracy, a scaled version of the Brier Score, was calculated as one minus the Brier Score of the iBox model divided by the Brier Score of the “null model”, obtained with the Kaplan-Meier estimator. Brier Scores’ 95% confidence intervals were computed following Blanche *et al*^27^.

The clinical utility was assessed with decision curves^28^ at 3-, 5-, and 7-years post-risk evaluation across risk thresholds ranging from 0% to 40%. The net benefit was calculated every 2.5% with a smoother (span of 0.1), and plotted along with the “treat all” and “treat none” reference strategies.

Performance metrics and plots were assessed in the whole study cohort, and in subgroups defined by race (“African American recipients” and “non-African American recipients”). A Z-test for independent samples was used to test for the statistical significance of differences observed between subgroups for key performance metrics^29^.

To compare the performances of the iBox between African American and non-African American kidney transplant recipients within the study cohort, heterogeneity of performances between subgroups was assessed using an interaction term between iBox score values and recipient’s race, in a Cox proportional hazards regression model predicting the outcome. A p-value <0.05 for the interaction term would indicate a statistically significant impact of recipient’s race on the iBox’s predictive performance. The analysis was performed both with and without stratification on the transplant center.

All tests were two-sided, and a p-value□<0.05 was considered significant. Continuous variables were reported as mean (standard deviation) or median (interquartile range), and categorical variables were reported as number (percentage).

#### Software

All analyses were performed using R (The R Foundation for Statistical Computing, Vienna, Austria).

#### Study reporting

Our manuscript complies with the relevant reporting guideline, namely the Transparent Reporting of a multivariable prediction model for Individual Prognosis or Diagnosis (TRIPOD+AI) statement^30^. The completed checklist is available in **Supplementary Methods 1**.

## RESULTS

### Characteristics of the cohort

The study included a total of 3,588 consecutive kidney transplant recipients originating from North American centers. Baseline characteristics are presented in **Table 1** and baseline characteristics stratified by transplant center are presented in **Supplementary Table 1**. The median time from transplantation to risk evaluation was 1.00 year (IQR 0.57-1.14). 481 (13.4%) experienced graft loss. The median follow-up time post-risk evaluation was 3.87 years (IQR 1.90 to 5.89).

**Table 1.** Baseline characteristics of the cohort, and according to the recipient’s race.

|  | n | Overall<br>N = 3,588<sup>1</sup> | Recipient's race |  | p-value |
| --- | --- | --- | --- | --- | --- |
|  |  |  | African American<br>n = 866<sup>1</sup> | Non-African<br>American<br>n = 2,722<sup>1</sup> |  |
| Recipient's characteristics |  |  |  |  |  |
| Age (years), mean (SD) | 3,585 | 49 (14) | 49 (13) | 50 (14) | 0.038 |
| Female gender, n (%) | 3,578 | 1,400 (39%) | 345 (40%) | 1,055 (39%) | 0.54 |
| Cause of kidney disease, n (%) | 2,727 |  |  |  | <0.001 |
| Diabetes |  | 597 (22%) | 147 (23%) | 450 (22%) |  |
| Glomerulonephritis |  | 754 (28%) | 130 (20%) | 624 (30%) |  |
| Vascular |  | 441 (16%) | 231 (36%) | 210 (10%) |  |
| Other |  | 935 (34%) | 139 (21%) | 796 (38%) |  |
| Donor's characteristics |  |  |  |  |  |
| Age (years), mean (SD) | 3,460 | 41 (15) | 39 (15) | 42 (14) | <0.001 |
| Female gender, n (%) | 3,467 | 1,760 (51%) | 406 (48%) | 1,354 (52%) | 0.058 |
| Hypertension, n (%) | 2,647 | 374 (14%) | 136 (21%) | 238 (12%) | <0.001 |
| Diabetes mellitus, n (%) | 2,368 | 85 (3.6%) | 42 (6.8%) | 43 (2.5%) | <0.001 |
| Deceased, n (%) | 3,588 | 1,797 (50%) | 628 (73%) | 1,169 (43%) | <0.001 |
| Cause of death, n (%) | 1,167 |  |  |  | <0.001 |
| Anoxia |  | 152 (13%) | 40 (9.3%) | 112 (15%) |  |
| Trauma |  | 223 (19%) | 62 (14%) | 161 (22%) |  |
| Vascular |  | 356 (31%) | 139 (32%) | 217 (29%) |  |
| Other |  | 436 (37%) | 190 (44%) | 246 (33%) |  |
| Extended criteria, n (%) | 2,063 | 295 (14%) | 106 (18%) | 189 (13%) | 0.003 |
| Transplantation characteristics |  |  |  |  |  |
| Prior kidney transplant, n (%) | 2,666 | 493 (18%) | 100 (16%) | 393 (19%) | 0.036 |
| Cold ischemia time (hours), mean (SD) | 2,623 | 10 (11) | 14 (12) | 8 (10) | <0.001 |
| Delayed graft function, n (%) | 3,378 | 463 (14%) | 207 (25%) | 256 (10.0%) | <0.001 |
| Number of HLA A/B/DR mismatches, mean (SD) | 2,615 | 3.6 (1.8) | 4.3 (1.4) | 3.5 (1.8) |  |
| iBox score evaluation |  |  |  |  |  |
| Time to risk evaluation (years), median (IQR) | 3,588 | 1.0 (0.6-1.1) | 1.0 (0.5-1.4) | 1.0 (0.6-1.1) |  |
| Estimated GFR (ml/min/1.73 m²), mean (SD) | 3,588 | 49 (22) | 47 (22) | 50 (22) | <0.001 |
| uPCR (g/g), median (IQR) | 3,588 | 0.1 (0.1-0.6) | 0.2 (0.1-1.1) | 0.1 (0.1-0.4) |  |
| MFI of the immunodominant DSA | 3,588 |  |  |  | 0.028 |
| < 1400 |  | 3,038 (85%) | 713 (82%) | 2,325 (85%) |  |
| ≥1400 |  | 550 (15%) | 153 (18%) | 397 (15%) |  |
| Protocol biopsy, n (%) | 1,102 | 208 (19%) | 30 (15%) | 178 (20%) | 0.17 |
| IFTA score | 3,588 |  |  |  | <0.001 |
| 0-1 |  | 3,054 (85%) | 693 (80%) | 2,361 (87%) |  |
| 2 |  | 390 (11%) | 126 (15%) | 264 (9.7%) |  |
| 3 |  | 144 (4.0%) | 47 (5.4%) | 97 (3.6%) |  |
| cg score | 3,588 |  |  |  | 0.50 |
| 0 |  | 3,289 (92%) | 789 (91%) | 2,500 (92%) |  |
| ≥1 |  | 299 (8.3%) | 77 (8.9%) | 222 (8.2%) |  |
| g+ptc score | 3,588 |  |  |  | <0.001 |
| 0-2 |  | 3,270 (91%) | 761 (88%) | 2,509 (92%) |  |
| 3-4 |  | 259 (7.2%) | 91 (11%) | 168 (6.2%) |  |
| 5-6 |  | 59 (1.6%) | 14 (1.6%) | 45 (1.7%) |  |
| i+t score | 3,588 |  |  |  | 0.68 |
| 0-2 |  | 3,126 (87%) | 751 (87%) | 2,375 (87%) |  |
| ≥3 |  | 462 (13%) | 115 (13%) | 347 (13%) |  |

<sup>1</sup>n (%); Mean (standard deviation) for normally distributed variables or median (interquartile range) for non-normally distributed variables
DSA, donor-specific antibody; eGFR, estimated glomerular filtration rate; HLA, human leukocyte antigen; MFI, mean fluorescence intensity; uPCR: urine protein-to-creatinine ratio.

African American kidney transplant recipients comprised 866 (24.1%) recipients of the study cohort. Compared to non-African American kidney transplant recipients, African American kidney transplant recipients more frequently experienced delayed graft function, had longer cold ischemia times and had higher numbers of HLA A/B/DR mismatches, and had higher uPCR and lower eGFR at the time of risk evaluation (**Table 1**).

### iBox evaluation in African American and non-African American kidney transplant recipients

African American KTRs had higher values of the iBox score, denoting a higher predicted probability of death-censored graft loss during follow-up (**Supplementary Figure 1**). Consistent with the higher iBox score, in the univariate survival analysis (**Table 2** and **Supplementary Figure 2**), African American KTRs had a higher hazard of experiencing death-censored graft loss at 7 years (HR: 2.14, 95% CI: 1.78 to 2.58, p<0.001). This difference was not explained by a longer time from transplant to evaluation, which was similar in both subgroups (1 year median in both groups; **Table 1**). At 3 and 5 years following risk evaluation, respectively 2,180 (60.8%) and 1,223 (34.1%) KTRs were still at risk (i.e., had not experienced the outcome and were still undergoing follow-up; **Supplementary Figure 2**).

**Table 2.** Univariate and multivariate Cox proportional hazards regression of time to death-censored graft loss. Patients were followed-up from the time of iBox score evaluation until graft loss, death, loss-to-follow-up or 7 years post-risk evaluation, whichever came first. Heterogeneity of performances between subgroups was assessed using an interaction term between iBox score values and recipient’s race, in a Cox proportional hazards regression model predicting death-censored graft loss. A p-value <0.05 for the interaction term would indicate a statistically significant impact of recipient’s race on the iBox’s predictive performance. The analysis was performed both without (left side) and with (right side) stratification on the transplant center.

|  |  | No stratification |  |  |  |  |  | Stratified by transplant center |  |  |  |  |  |
| --- | --- | --- | --- | --- | --- | --- | --- | --- | --- | --- | --- | --- | --- |
|  |  | Univariate |  | Multivariate,<br>without interaction |  | Multivariate,<br>with interaction |  | Univariate |  | Multivariate,<br>without interaction |  | Multivariate,<br>with interaction |  |
| Variable | No. of patients<br>(No. of events) | Hazard ratio<br>(95% CI) | p-value | Hazard ratio<br>(95% CI) | p-value | Hazard ratio<br>(95% CI) | p-value | Hazard ratio<br>(95% CI) | p-value | Hazard ratio<br>(95% CI) | p-value | Hazard ratio<br>(95% CI) | p-value |
| **iBox score** | 3,588<br>(481) | 2.47<br>(2.31;2.64) | **< 0.001** | 2.45<br>(2.29;2.62) | **< 0.001** | 2.45<br>(2.26;2.65) | **< 0.001** | 2.42<br>(2.26;2.60) | **< 0.001** | 2.40<br>(2.24;2.58) | **< 0.001** | 2.42<br>(2.22;2.64) | **< 0.001** |
| **Race** |  |  |  |  |  |  |  |  |  |  |  |  |  |
| **Non-African American**<br>**(reference group)** | 2,722<br>(304) | 1.00 | - | 1.00 | - | 1.00 | - | 1.00 | - | 1.00 | - | 1.00 | - |
| **African American** | 866<br>(177) | 2.14<br>(1.78;2.58) | **< 0.001** | 1.16<br>(0.96;1.40) | 0.12 | 1.17<br>(0.85;1.61) | 0.35 | 1.83<br>(1.49;2.24) | **< 0.001** | 1.21<br>(0.98;1.49) | 0.08 | 1.25<br>(0.90;1.74) | 0.19 |
| **Interaction between**<br>**iBox and race** | 3,588<br>(481) | - | - | - | - | 1.00<br>(0.87;1.15) | 0.97 | - | - | - | - | 0.98<br>(0.85;1.13) | 0.80 |

CI: confidence interval

In a Cox proportional hazards model, no significant interaction was found between the iBox score value and the recipient’s race, in both the unstratified analysis (p=0.97) and the analysis stratified by transplant center (p=0.80) (**Table 2**).

### Predictive performance of the iBox in African American and non-African American kidney transplant recipients

In the study cohort and in both subgroups defined by recipients’ race, the iBox achieved good discrimination, with a c-index of 0.84 (95% CI: 0.82-0.85) in the study cohort, 0.81 (95% CI: 0.78-0.84) in African American KTRs, and 0.83 (95% CI: 0.81-0.85) in non-African American KTRs (**Table 3**). No significant difference was found between the c-index of the African American KTRs and the non-African American KTRs (p=0.25).

**Table 3.**
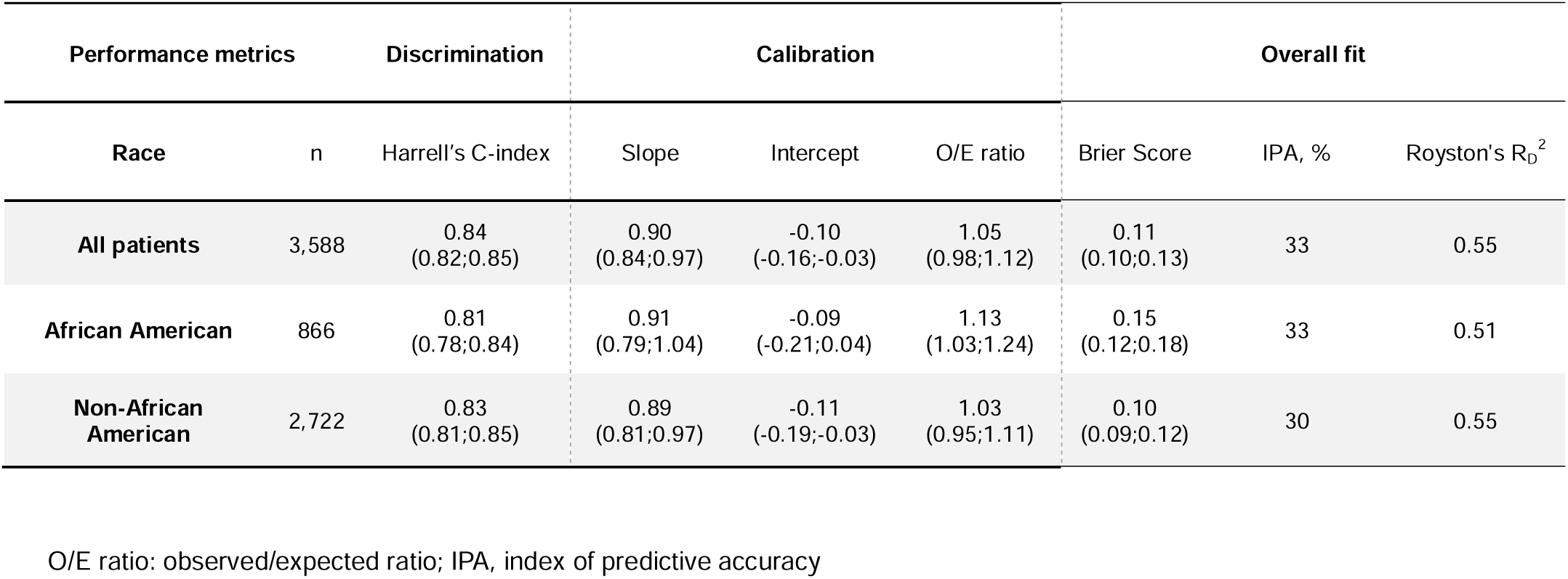
Predictive performance of the iBox prognostication system in the study cohort and in African American and non-African American recipients. Discrimination, calibration and overall accuracy performance metrics are presented across all time points post-risk evaluation up to 7 years in the study cohort, in African American, and in non-African American.

In terms of calibration, the agreement between the iBox’s predicted and observed risks was adequate in the study cohort and in both the African American and non-African American recipients, as reflected in the calibration metrics (calibration slope 0.90 [95%CI: 0.84-0.97], 0.91 [95%CI: 0.79-1.04], and 0.89 [95%CI: 0.81-0.97] for the study cohort, African American recipients, and non-African American recipients, respectively; **Table 3**) and the calibration plot at 3-, 5- and 7-years post-risk evaluation (**Figure 1**). The observed/expected ratio was equal to 1.05 (95% CI: 0.98-1.12) in the study cohort, 1.13 (95%CI: 1.03-1.24) in African American KTRs, and 1.03 (95%CI: 0.95-1.11) in non-African American KTRs. No significant difference was found between the calibration slopes, intercepts, and observed/expected ratios of the African American KTRs and the non-African American KTRs (p=0.75, p=0.75, and p=0.13 for the calibration slope, intercept, and observed/expected ratio, respectively).

**Figure 1.**
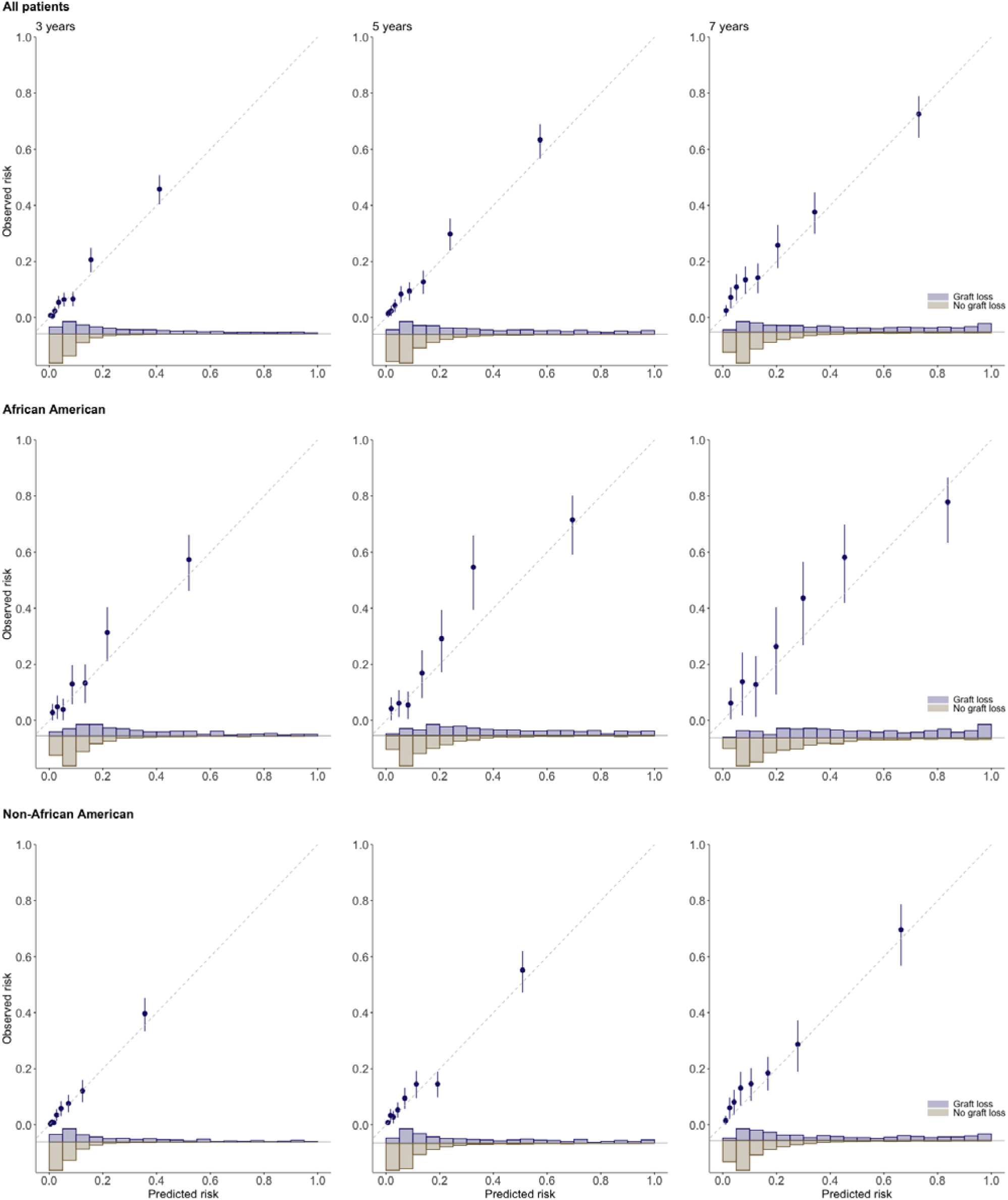
Calibration assessed at 3-, 5- and 7-years following iBox evaluation in the study cohort and in African American and non-African American recipients. Calibration plots are presented at 3-, 5- and 7-years post risk-evaluation in the study cohort (top panel), in African American (middle panel), and in non-African American (lower panel). In each group, the median of the predicted risks was plotted against the observed event probability estimated by (one minus) the Kaplan-Meier estimator. The diagonal line at the origin represents the perfectly calibrated model. The histograms represent the distribution of the model’ individual predicted risks according to the event status.

Regarding overall fit, Brier score values indicated appropriate accuracy, with 0.11 (95% CI: 0.10-0.13) in the study cohort, 0.15 (95% CI: 0.12-0.18) in African American KTRs, and 0.10 (95% CI: 0.09-0.12) in non-African American KTRs (**Table 3**). A significant difference was found between the Brier Scores of the African American KTRs and the non-African American KTRs (p=0.007).

In the decision curve analysis, the strategy “treat per iBox” led to the largest net benefit across the range of threshold probabilities, in comparison to the “treat all” and “treat none” strategies, in the study cohort and in African American KTRs at 3-, 5- and 7-years post-risk evaluation (**Supplementary Figure 3**).

### Predictive performance of the iBox in African American and non-African American kidney transplant recipients across different eGFR equations

When the eGFR was estimated with the MDRD_186_ equation or the KRS equation, the discrimination and overall fit yielded very similar results, with a c-index of 0.83 (95% CI: 0.82-0.85) in the study cohort, 0.81 (95% CI: 0.78-0.84) in African American KTRs, and 0.83 (95% CI: 0.81-0.85) in non-African American KTRs for the MDRD_186_ equation (**Supplementary Table 2**), and a c-index of 0.84 (95% CI: 0.82;0.85) in the study cohort, 0.81 (95% CI: 0.78;0.84) in African American KTRs, and 0.83 (95% CI: 0.81;0.85) in non-African American KTRs for the KRS equation (**Supplementary Table 3**).

However, when using the MDRD_186_ equation, the calibration metrics were slighlty poorer in African American KTRs (calibration slope 0.88 [95% CI: 0.76-1.00] in African American KTRs; **Supplementary Table 2**), whereas when using the KRS equation, the calibration slope and intercept were slightly improved (calibration slope 0.94 [95% CI: 0.82-1.07] in African American KTRs; **Supplementary Table 3**). Calibration plots for both equations can be found in **Supplementary Figures 4 and 5**.

### Predictive performance of the iBox across diverse subpopulations

We investigated the prediction performance of the iBox when applied in a series of distinct subpopulations, including living and deceased donors, according to recipient age and sex, and donor type (extended or standard criteria). The iBox discriminative capability was accurate in these scenarios (c-index range 0.80 to 0.85; **Supplementary Table 4**).

When stratifying by race, the iBox maintained good discrimination in both subgroups across these scenarios (c-index range 0.78 to 0.85 in African American recipients, c-index range 0.81 to 0.85 in non-African American recipients; **Supplementary Table 5**). The low number of patients in the scenarios in the African American subgroup did not allow for an assessment of calibration.

## DISCUSSION

In this multicenter evaluation study performed in 3,588 North American KTRs, the iBox was accurate in predicting graft loss up to 7 years following evaluation, in terms of overall fit, calibration, discriminative ability, and demonstrated clinical utility. The iBox’s performance in African Americans was similar to that of non-African American recipients.

Our findings indicate a marginal underestimation of the true risk of graft loss in African American KTRs. However, performance metrics indicated an appropriate performance overall and did not appear to differ between groups. Altogether, our results support that the iBox prognostication system can be used, in routine care or in clinical trials, to generate accurate predictions in African American recipients.

In univariate analysis, a significantly greater risk of graft loss was observed in the African American population. Consistent with these data, the OPTN/SRTR 2022 Annual Data Report describes lower 5-year graft survival in African American KTRs for both living and deceased donor transplants^31^. This difference was no longer observed in the multivariable analysis stratifying by study center and adjusting using the iBox score value. Whether this denotes a true phenomenon (i.e., no independent effect of race on outcomes) or is due to residual confounding not accounted for in this study requires further research. Regardless, it stresses the importance of carefully accounting for confounding in causal inference studies in the field of kidney transplantation, especially when studying associations between self-declared race and outcomes. In this regard, relying on a validated risk score (e.g., the iBox) appears promising. In such studies, it is equally important to account for confounding factors that may arise from socioeconomic or genetic differences^14^ between African American and non-African American kidney transplant recipients.

The iBox may be an important tool to assist in individual patient management, but also to accelerate the development of new therapeutics, where superiority could be demonstrated on a surrogate marker that is prognostic and predictive of long-term graft loss.

Our study opens the door to exploring performance with a complete breakdown by race. However, comparing a model’s performance across more than two groups comes with methodological challenges, as each group needs a sufficient sample size and number of events while still including consecutive KTRs. It would also require more advanced modeling strategies to properly handle the multi-group design (e.g., using multinomial regression instead of logistic regression).

Our study includes the largest cohort of African American kidney transplant recipients, where the performance of the iBox was assessed, and is key to further confirming the generalizability of the iBox qualification data^8^ to the African American kidney transplant population, who are in great need of therapies that improve long-term outcomes and are frequently underrepresented in clinical trials^32^. In addition, selection bias appears to be controlled in our study, given the inclusion of consecutive KTRs within each center and the percentage of African American KTRs in the cohort (24.1%), which aligns with what is observed in the general population of KTRs, as reported in the OPTN/SRTR Annual Report^31^. Our study confirms the fairness of the iBox, meaning that its use as an outcome measure in a clinical trial will not lead to wrongfully restricting the use of a drug in African American KTRs.

Although exploratory by nature, results from the subgroup analysis suggest that performance is adequate in both African American and non-African American recipients across subgroups defined by the recipient’s age and gender, donor’s living status and type, and biopsy indication. In addition, the stability of beta-coefficients in the unstratified and stratified Cox models indicates limited between-centers heterogeneity regarding performance, thereby strengthening the generalizability of our findings.

Although era-specific variations in practices and outcomes (e.g., the COVID-19 pandemic) should be considered when interpreting our results, they are unlikely to significantly affect our findings. First, era-specific variations overlap with center-specific variations, as each cohort has a distinct recruitment period. However, we observe consistency in performance across centers, both in the overall population and among African American and non-African American recipients. Second, in the original iBox publication^4^, it has been demonstrated that the iBox effectively predicts outcomes regardless of variations in practices, as evidenced for instance, by its appropriate performance in the Certitem (NCT01079143), Borteject (NCT01873157), or Rituxerah (EudraCT 2007-003213-13) trials. Further work is ongoing to assess the stability of the iBox’s performance over time.

We acknowledge that the collection of the race variable was performed heterogeneously across centers, not providing us with sufficient granularity to evaluate the iBox in other subpopulations (e.g., Hispanic or Asian recipients). In addition, we used self-declared race, as it is recommended by AJT’s editorial guidelines and the American Medical Association^33^. While this approach is not perfect, its potential limitations are likely of no consequence for the present study, as classification regarding African American race was available across all centers, enabling us to reliably distinguish between African American and non-African American recipients, and, consequently, to accurately assess the iBox’s performance in this particular population. Furthermore, the use of the center as a stratification variable in the analyses mitigates the impact of between-centers variations with regard to the way race was collected. We also acknowledge that we did not include African American KTRs from the southeastern part of the US. To what extent this leads to precautions with regard to the generalizability of our results in this specific population is unknown. We acknowledge that outcome capture using OPTN data might be incomplete^34,35^. However, this is likely not differential and is mitigated by the fact that outcomes were ascertained in our study by local investigators relying on multiple sources (e.g., institutional EHRs) rather than solely on OPTN data.

## CONCLUSION

This multicenter study shows that the iBox prognostication system predicts graft loss with similar accuracy in African American and non-African American kidney transplant recipients, supporting fairness in its use as an efficacy endpoint in clinical trials.

## Ethical approval

The protocol of this study was approved by the Institutional Review Board of the Paris Transplant Institute. Data were sent anonymized to the Paris Transplant Group. The institutional review boards of the Paris Transplant Group participating centres approved the study. This research is governed by the CNIL (French Data Protection Agency) “Reference Methodology for processing personal data used with the scope of health research” (amended MR-001). The study was conducted in accordance with the Declaration of Helsinki and the International Conference on Harmonization Guidelines for Good Clinical Practice, and the Declaration of Istanbul on Organ Trafficking and Transplant Tourism.

## Data and code sharing

The datasets and analytical codes used in this study are available from the corresponding author on reasonable request, subject to relevant institutional and ethical approvals. The French company Okeiro develops a software solution to support the deployment and clinical implementation of the iBox system in transplant centers worldwide.

## Protocol and registration

iBox prognostication system (https://clinicaltrials.gov, NCT03474003).

## Patient and public involvement

No involvement.

